# The evaluation of a web‐based tool for measuring the uncorrected visual acuity and refractive error in keratoconus eyes: a prospective open‐label method comparison study

**DOI:** 10.1101/2021.03.01.21252664

**Authors:** Marc B Muijzer, Janneau L J Claessens, Francesco Cassano, Daniel A Godefrooij, Yves F D M Prevoo, Robert P L Wisse

## Abstract

**Purpose:** To evaluate the outcome of a web‐based digital assessment of visual acuity and refractive error, compared to a manifest refraction assessment, in keratoconus patients with complex refractive errors.

**Material and methods:** Keratoconus patients, aged 18 to 40, with a refractive error between ‐6 and +4 diopters were eligible. Each participant subsequently underwent an uncorrected visual acuity and a refractive assessment. Refractive error was assessed with the web‐based tool (index test), an autorefractor, and a manifest refraction (reference test) by an optometrist. Corrected visual acuity was assessed with the web‐based and manifest refractive prescription. Non‐inferiority was defined as the 95% limits‐of‐agreement (95%LoA) of the differences in spherical equivalent between the index and reference test not exceeding +/‐ 0.5 diopters. Agreement was assessed by an intraclass correlation coefficient and Bland‐Altman analyses.

**Results:** A total of 100 eyes of 50 patients were examined. The overall mean difference of the uncorrected visual acuity measured ‐0.01 LogMAR (95%LoA:‐0.63–0.60). The variability decreased in the high uncorrected visual acuity subgroup (mean difference: 0.15 LogMAR, 95%LoA:‐0.25–0.55). The intraclass correlation coefficient of the three refractive assessments was 0.32. The overall mean difference in spherical equivalent between the index and reference test measured ‐0.58 diopters (95%LoA:‐4.49 – 3.33, P=0.008). The mean differences for myopic and hyperopic subjects were 0.09 diopters (P=0.675) and ‐2.06 diopters (P<0.001), respectively.

**Conclusions:** Our results show promising results in the ability of the web‐based tool to remotely assess visual acuity in keratoconus patients. The agreement is better in higher visual acuity ranges and could provide physicians with an objective measurement to enhance teleconsultations, especially relevant when access to health care is limited. The assessment of the refractive error using the web‐based tool was found to be inferior to the manifest refraction and deserves further training of the tools algorithm.

## Introduction

Globally, an estimated 1 billion people have a visual impairment that can be prevented or is undetected and this number is expected to rise.(1,2) Main causes of visual impairment are refractive errors, cataract or chronic ophthalmic conditions (e.g. macular degeneration). In an aging population, the demand for eye care is increasing rapidly, which imposes a challenge for providers of eye care in both developed and less‐developed countries.(1,3) The acutely reduced access to healthcare during the COVID‐19 outbreak underlined the need for a paradigm shift in the delivery of eye care.(4)

To become less reliant on hospital facilities and trained professionals for monitoring eye conditions, there is a growing interest in telemedicine and digital tools. In particular, promising advances have been made for automatic assessment of retinal images.(5) The refractive error and visual acuity are considered important clinical parameters for diagnosing and monitoring eye conditions and as such various tools to remotely assess refractive errors and visual acuity are developed and clinically validated.(6,7)

Currently, the manifest refraction – consisting of the measurement of the refractive error and the visual acuity – by a trained professional is considered the gold standard.(8) Automated assessment of the refractive error using an autorefractor is considered non‐inferior to the manifest refraction in healthy eyes.(9,10) However, the reliability of the autorefractor decreases in eyes suffering from complex refractive errors (a hallmark sign of keratoconus) and it does not measure visual acuity.(11) Moreover, both the manifest and automated techniques require (expensive) medical equipment and/or qualified personnel and makes them unsuitable for home monitoring or use in less‐developed countries.

Recently the authors published the outcomes of the “manifest versus online refraction evaluation”‐ trial, which reports the validation of a web‐based refractive assessment in healthy adults. The tool was found non‐inferior to a manifest refraction performed by a trained optometrist and is accessible via https://easee.online.(6) Notably, the tool does not require any specialized equipment – it only requires a mobile phone and a computer screen – and thus can be performed in most home environments. However, the authors acknowledge that the results within a healthy population are not necessarily representative for individuals with a suboptimal visual performance. Therefore the design of the MORE‐trial included a cohort of keratoconus patients to evaluate the outcome of the digital refraction tool in a population with an eye condition. Keratoconus was chosen as these patients are often still able to achieve an acceptable visual acuity with a proper prescription, despite their complex refractive errors, including irregular astigmatism. (11,12) Keratoconus is an uncommon condition typically diagnosed in adolescents, where gradual thinning of the cornea leads to a progressive ectasia and subsequent irregular astigmatism.(13,14) With its insidious onset, many early patients are unaware of this diagnosis, and keratoconus patients therefore pose a challenge for any assessment of refractive error.(11)

Here, we present the results of the keratoconus cohort of the MORE trial, a validation study of a web‐based tool for the assessment of visual acuity and refractive error.

## Material and methods

### Study Design and Recruitment

Data were prospectively collected in the open‐label single‐center method comparison “manifest versus online refraction evaluation”‐trial, performed at the University Medical Center Utrecht in Utrecht, the Netherlands. Participants enrolled into the study were patients who visited the outpatient clinic at the University Medical Center Utrecht in Utrecht. Inclusion criteria were an established diagnosis of keratoconus, a clear central cornea, and an age between 18 to 40 years. Subjects were excluded if their manifest refractive error, converted to spherical equivalent, was worse than ‐6 diopter (D; for myopia) or +4 D (for hyperopia). Furthermore, we excluded subjects who had undergone corneal crosslinking 6 months prior to study participation to diminish effects of corneal haze, who had diabetes, who were pregnant or lactated, or who were unable to perform the a web‐based refraction assessment (Figure 1). All participants provided written informed consent and were enrolled in the study between January 31, 2018 and June 23, 2019.

**Figure 1.**
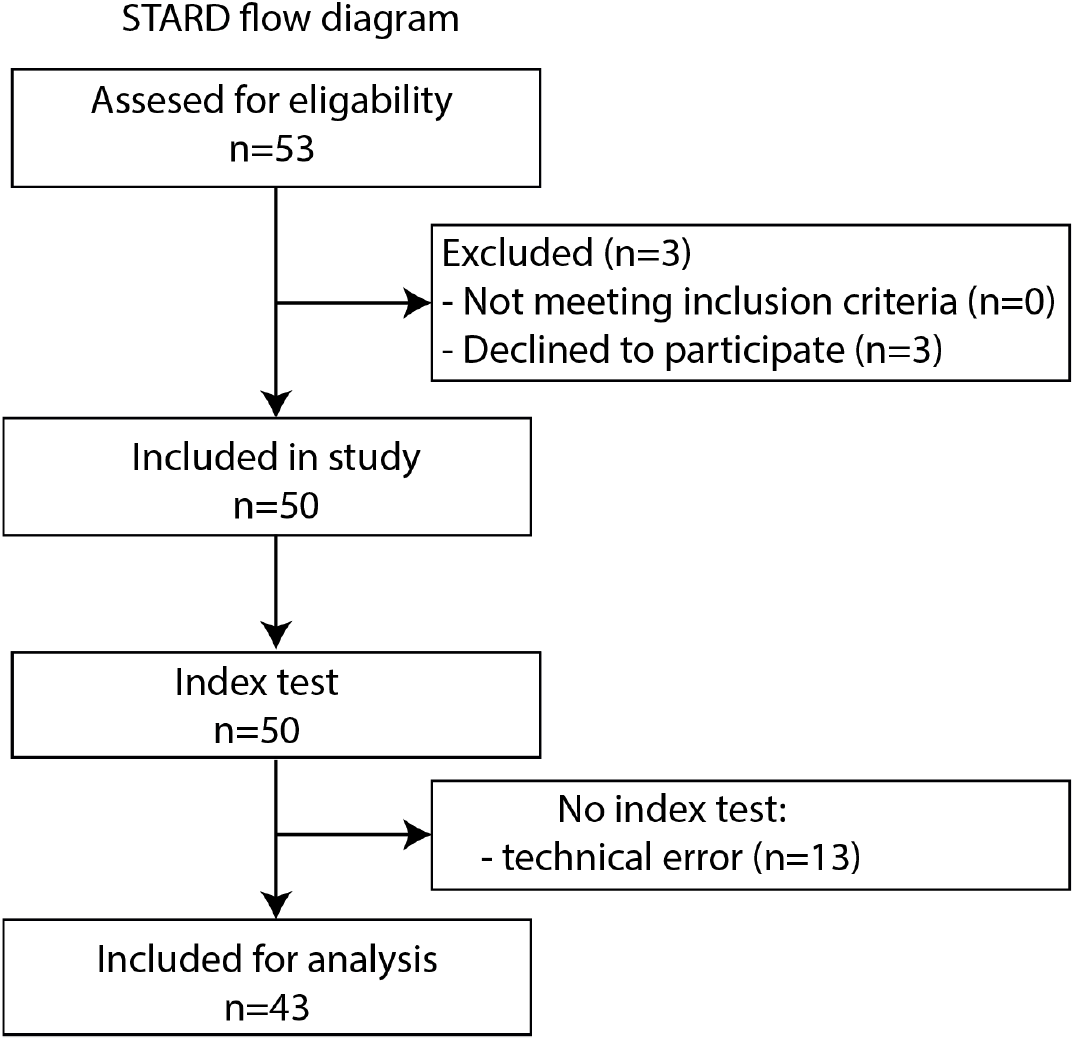
STARD flow diagram for MORE trial. STARD flow showing participant flow through each stage om the open label method comparison trial.

All subjects underwent 3 consecutive test designed to determine the refractive state of both eyes in the following order and the subject was blinded for the outcome of all tests. First, the refractive error was measured using autorefraction (Topcon RM 8800, Topcon Corporation, Japan) and corneal imaging was performed using Scheimpflug tomography (Pencatam HR, Oculus GmbH, Germany).

Second, an optometrist performed the reference test (manifest subjective refraction). Finally, the subject performed the index test using the digital refraction tool. The digital refraction tool is web‐based and was a custom version of the commercially available Easee refractive assessment tool, specifically built for this clinical trial and is identical to the second‐generation algorithm we have described previously.(6) In short, a smartphone functions as a remote control by which the user submits input from a distance of 3 meters to a computer screen that displays the Web‐based assessment. The user is presented a sequence of optotypes and astigmatism dials. Any visual acuity below 1.0 (worse than 20/20) is considered to be caused by a refractive error. The web‐based refraction assessment is classified as a *Conformité Européenne* class 1m medical device, which is in accordance with European Union Medical Device Regulation 2017/745, and the software is classified as class A, which is in accordance with International Electro technical Commission standard 62304:2014.

The uncorrected distance visual acuity (UDVA) was recorded using an Early Treatment Diabetic Retinopathy Study (ETDRS) visual acuity wallchart and the web‐based visual acuity test. Corrected distance visual acuity (CDVA) was measured using a correction based on the results of the manifest and web‐based refraction assessment outcome. Visual acuity was tested in accordance with International Organization for Standardization standard 8596, with regard to optotypes and room illumination.(15) The projected optotypes were randomized to mitigate any possible test‐retest effect. The study protocol did not cover the assessment of CDVA with the autorefraction result, as previous research has shown a low reliability of autorefraction measurement in keratoconus eyes.(11)

The following data were recorded for each participant/eye: age, gender, laterality, (ophthalmic) medical history, Amsler‐Krumeich stage (AK1: mild, AK2: moderate, AK3: severe), mean & maximum keratometry, previous prescription (if known), use of spectacles or contact lenses, UDVA, CDVA, and refractive outcome, including spherical and cylindrical power (in D) and axis (in degrees).

All procedures were performed in accordance with the Declaration of Helsinki, local and national laws regarding research (i.e. the Act on Scientific Research Involving Humans), European directives with respect to privacy (General Data Protection Regulation 2016/679) and medical devices (Medical Device Regulation 2017/745), and the 2015 Standards for Reporting Diagnostic Accuracy Studies.(16) The study protocol was approved by our institution’s Ethics Review Board (METC number: 17‐524), and it was registered at clinicaltrials.gov (number: NCT03313921) and CCMO (number: NL61478.041.17).

### Statistical analysis

The primary study outcome was refractive error as measured using the web‐based refractive assessment and compared with the subjective manifest refraction and autorefraction. The signation of the refractive error (+/‐), spherical power, cylindrical power were converted into a spherical equivalent. The agreement of the spherical equivalent of the various methods was compared using a two‐way mixed effect intraclass correlation coefficient (ICC) of a single measurement. In addition, the difference between the measurements of the web‐based and manifest assessment were compared using a Fourier analysis.(17,18) Specifically, we analyzed the signation of the refractive error (+/‐), spherical power, cylindrical power, and axis, which were converted into power vectors. Subsequently, the difference between the power vectors of the various methods was calculated as a residual vector (i.e. a vector of the difference). The power vectors are non‐linear in nature, which precludes statistical analysis of differences between power vectors. Secondary study outcomes included the UDVA and CDVA, the latter measured using the outcome of both the manifest and web‐based refractive assessment. A post‐hoc subgroup analysis was performed for high and low vision visual acuity ranges (high: <=0.5 logMAR, low >0.5 LogMAR), with a cut‐off as defined by the World Health Organization.(19)

The data was assessed for normality of the distribution. Groups were compared using the two‐tailed paired Student’s *t* test. In addition, a multivariable generalized estimates equation (GEE) analysis of the difference between the power vectors, was used to correct for bilaterality (both eyes of the same patient included), keratoconus severity, age, and sex. For keratoconus severity, Amsler‐Krumeich stages 2 and 3 were combined, due to the low number of cases. Missing cases were not included in the analysis, nor imputed, since these were not considered *missing at random*. Differences with a *P* value <0.05 were considered statistically significant. The outcomes were stratified for myopia, hyperopia, and keratoconus stages. The non‐inferiority limit was set at 0.5 D to reflect a clinically significant difference in line with literature.(20) Non‐inferiority was defined as the 95% limits of agreement of the difference in spherical equivalent between the web‐based refractive assessment and the manifest refraction not exceeding +/‐ 0.5. The currently applied method (comparing limits of agreements rather than ICC) is more stringent and better in line with up‐to‐date insights in method comparison studies.(21)

The power calculation for the MORE trial was based on an intraclass correlation for the 3 different refraction methods, using the following formula in R: Sample size (p=0.70,p0=0,k=3,alpha=(0.05/12),tails=2, power=0.80, by=“p”, step=0.025). Data were analyzed using SPSS version 25.0 (IBM, Armonk, New York, USA) and R statistical software version 4.0.3 (CRAN, Vienna, Austria). For the Bland‐Altman analysis the “BlandAltmanLeh” (version 0.3.1) package was used.

## Results

A total of 100 eyes from 50 keratoconus patients were included in the study, no subjects were excluded. The clinical characteristics of the study population are summarized in Table 1 and all relevant variables are stratified for Amsler‐Krumeich keratoconus stage.(13) The majority of the participants was male (78%, n=39), and used visual aids to correct their refractive error (78%, n=39). Both results were expected; keratoconus is more prevalent in males and the hallmark cone shape in keratoconus induces myopia and (irregular) astigmatism.(13) A total of 23 underwent corneal crosslinking (n= 37 eyes), all resolved without sequelae. A total of 9 subjects reported ocular complaints at the time of the measurement; all subjects reported blurred vision. No adverse events or complications were recorded during the trial. The refractive error and visual acuity data was missing in 13 subjects (11 myopic and 2 hyperopic) because of a technical error in the web‐based refractive assessment.

**Table 1.** Clinical characteristics of the study population (100 eyes of 50 patients)

|  |  |  |  |  |
| --- | --- | --- | --- | --- |
| Age (years), mean ± SD |  | 25.6 ± 5.4 |  |  |
| Sex (male), n (%) |  | 39 (78) |  |  |
| Current use of visual aids, n (%) |  | 38 (78) |  |  |
| Spectacles, n (%) |  | 28 (56) |  |  |
| Contact lenses, n (%) |  | 17 (34) |  |  |
| Ocular complaints, n (%) |  | 9 (18) |  |  |
| Medication use, n (%) |  | 10 (20) |  |  |
| Previous surgical treatment, n (%) |  | 24 (48) |  |  |
| Distribution of refractive errors and keratoconus severity classification<sup>a</sup> |  |  |  |  |
|  | Total (n=100) | Mild keratoconus<br>AK stage 1<br>(n=84) | Moderate<br>keratoconus<br>AK stage 2<br>(n=13) | Severe keratoconus<br>AK stage 3 (n=3) |
| Mean keratometry, mean ± SD | 45.66 ±2.90 | 45.27 ±2.79 | 47.46 ±1.75 | 48.63 ±5.37 |
| Maximum keratometry, mean ± SD | 52.55 ±6.39 | 52.08 ±6.30 | 54.50 ±4.48 | 57.33 ±13.68 |
| Refractive error<sup>1</sup> | 100 | 84 | 13 | 3 |
| Hyperopia, n | 29 | 23 | 5 | 1 |
| Emmetropia, n | 5 | 5 | 0 | 0 |
| Mild myopia, n | 44 | 40 | 4 | 0 |
| Severe myopia | 22 | 16 | 4 | 2 |

Abbreviations: AK; Amsler-Krumeich stage, SD; standard deviation
<sup>a</sup> Mild myopia was defined as refractive error of $-3$ diopter or less; severe myopia was defined as refractive error worse than $-3$ diopter. Refractive error was determined on the basis of the spherical equivalent of the manifest refraction value, (reference test), and is reported for both eyes separately. The distribution of refractive errors was not significantly different between the classifications ( $P=0.30$ ).

### Intraclass Correlation Coefficients

The concordance of the refractive error among the three methods was assessed using the ICC of the spherical equivalent. The overall ICC of all 3 measurements was 0.32 (95% CI 0.19 ‐ 0.46). The ICC for the manifest refraction and web‐based refractive assessment was overall 0.36 (95% CI 0.22 ‐ 0.53) and for the mild keratoconus subgroup 0.48 (95% CI 0.29 ‐ 0.64). All eyes were included in the ICC because of the asymmetrical manifestation of keratoconus. ICC calculations for either all right or left eyes separately did not lead to new insights.

### Digital visual acuity testing

UDVA was measured using the digital index test and with an ETDRS visual acuity wall chart as reference. Mean UDVA measured digitally was logMAR 0.57±0.39 (Snellen 0.38±0.33), and with the ETDRS wall chart logMAR 0.58±0.52 (Snellen 0.46±0.40). The mean difference between the measurements was considered small and non‐significant (‐0.01 LogMAR, P=0.76). There was a considerable distribution in measurement differences (95% LoA: ‐0.63 – 0.60). In the high visual acuity subgroup (<=0.5 LogMAR) this variability decreased (mean difference: 0.15 LogMAR, 95% LoA: ‐0.25 – 0.55). In the low visual acuity subgroup the mean difference between both measurements was ‐0.20 LogMAR(95% LoA: ‐0.82 – 0.41). Figure 2 depicts the correlation of the web‐based visual acuity assessment compared with the ETDRS measurement in a Bland‐Altman plot. The visual acuity assessments do not agree equally through the range of measurements. As can be observed in the high visual acuity subgroup, the majority of the measures show little difference and the mean difference is mostly attributed to outliers. In the lower visual acuity ranges, the measurements appear to be much more variable.

**Figure 2.**
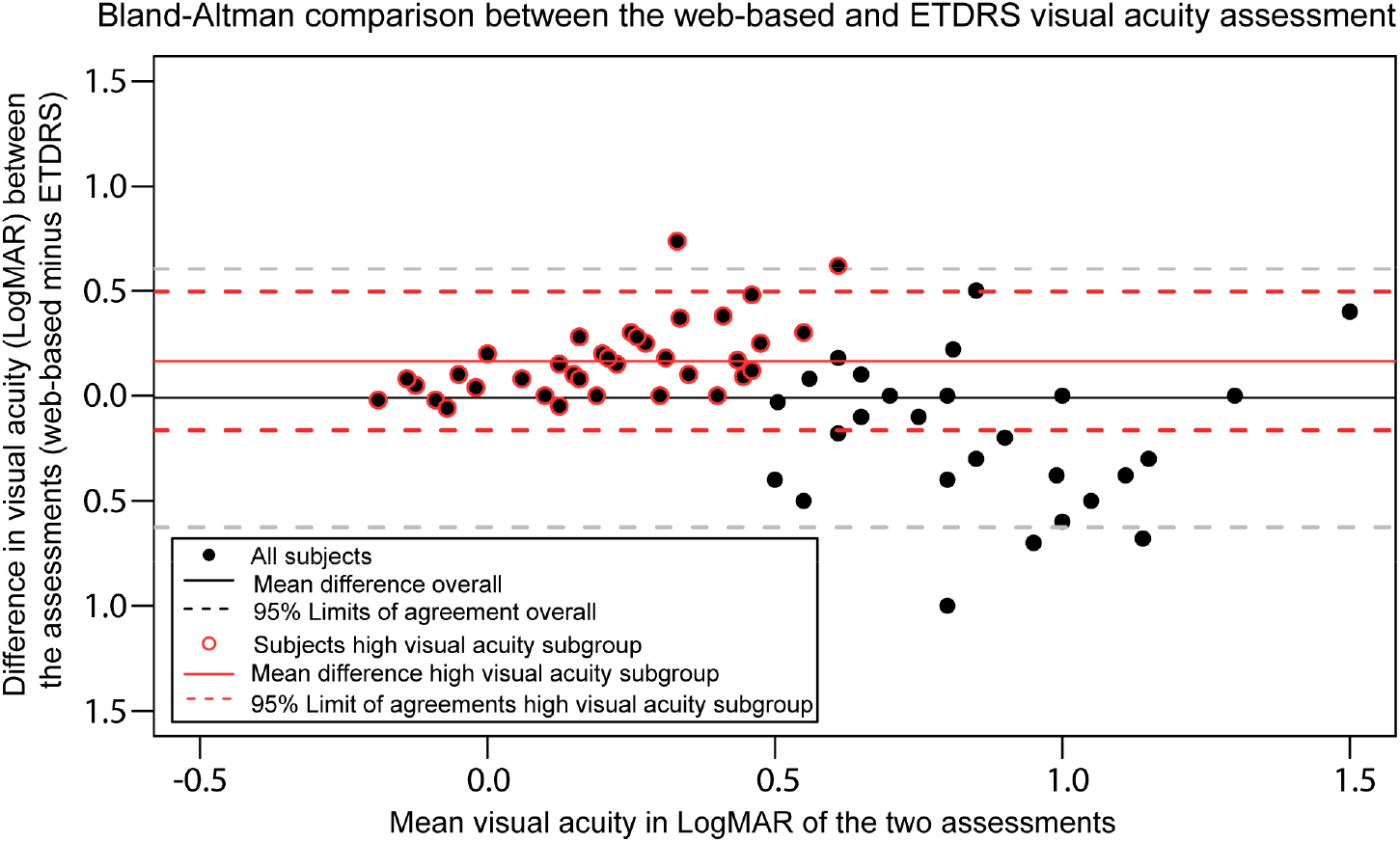
A Bland‐Altman plot displaying the differences in logarithmic minimum angle of resolution (LogMAR) between the web‐based uncorrected distance visual acuity assessment (index test) and the ETDRS uncorrected distance visual acuity measurement (reference test). The differences between the reference test and index test shown on the Y‐axis are expressed as the difference of the web‐based uncorrected distance visual acuity assessment outcome minus the ETDRS uncorrected distance visual acuity outcome. The x‐axis shows the mean visual acuity in LogMAR of the two assessments, where a more negative value represents a higher visual acuity. The outcome is stratified for a high visual acuity subgroup (uncorrected distance visual acuity <= 0.5 LogMAR) highlighted with a red circle.

### Overall outcome of the web‐based refraction assessment

Assessments of refractive error are reported stratified for myopia and hyperopia (see Table 2). Detailed outcomes per keratoconus stage are found in the supplementary tables (supplementary Table 1 and 2). Overall, the spherical equivalent of the refractive error measured using the manifest refraction and the web‐based refraction assessment differed significantly for both myopic (0.09D) and hyperopic subjects (‐2.06D). Albeit the relatively small mean difference, the 95% LoAof the difference of the spherical equivalent extended beyond the *a priori* set non‐inferiority limit of 0.5 D in both myopic subjects (95% LoA‐3.28 – 3.47) and hyperopic subjects (95% LoA ‐5.52 – 1.39). When transposed into power vectors these differences measure ‐1.08D for myopic subjects and ‐0.69D for hyperopic subjects (Table 2, top row). Figure 3 depicts the correlation of the web‐based refraction assessment compared with the reference test. The visualization shows a wide distribution of differences between the assessments overall. However, within the emmetropic/myopic refractive error group the agreement between the methods improves in the low refractive error ranges. Furthermore, it can be observed that the web‐based refractive assessment yields a more hyperopic outcome in higher myopes (i.e. undercorrected) and a more myopic refractive outcome in low myopics (i.e. overcorrected). When outcomes of the refractive assessment are broken down per keratoconus stage, the more advanced keratoconus cases (AK stage 2 and 3) show an increased difference between the index and reference test (supplementary data Table 1 and 2).

**Table 2.** Measured refractive error and visual acuity.

| Table 2. Measured refractive error and visual acuity |  |  |  |  |  |  |  |  |  |  |
| --- | --- | --- | --- | --- | --- | --- | --- | --- | --- | --- |
|  | Emmetropic and myopic subject |  |  |  |  | Hyperopic participants |  |  |  |  |
| Refractive error and visual acuity | Web-based refraction<sup>a</sup> (n=60) | Manifest Refraction<sup>a</sup> (n=71) | Difference<sup>b,c</sup> | 95% CI | P-value<sup>d</sup> | Web-based refraction<sup>a</sup> (n=27) | Manifest refraction<sup>a</sup> (n=29) | Difference<sup>b,c</sup> | 95% CI | P-value<sup>d</sup> |
| Power vector<sup>e,f</sup> (D) | 2.34<br>±1.02 | 2.69<br>±1.74 | -1.08 | -1.41 – -0.74 | N.A. | 1.35<br>±0.68 | 2.02<br>±1.04 | -0.69 | -0.92 – -0.46 | N.A. |
| Power vector J0<sup>f</sup> (X) | -0.08<br>±0.54 | -0.61<br>±1.17 | -1.16 | -1.41 – -0.90 | N.A. | -0.08<br>±0.26 | -1.27<br>±1.01 | -1.47 | -2.00 – -0.93 | N.A. |
| Power vector J45<sup>f</sup> (Y) | 0.05<br>±0.62 | 0.02<br>±0.86 | -0.72 | -0.91 – -0.53 | N.A. | -0.04<br>±0.23 | -0.16<br>±1.00 | -0.98 | -1.41 – -0.54 | N.A. |
| SEQ (D) | -1.96<br>±1.20 | -2.05<br>±1.76 | 0.09 | -0.35 – 0.54 | 0.675 | -1.02<br>±1.40 | 1.04<br>±0.83 | -2.06 | -2.76 – -1.37 | <0.001 |
| Sphere | -1.53<br>±1.15 | -0.87<br>±1.79 | -0.76 | -1.23 – 0.28 | N.A. | -0.59<br>±1.39 | 2.68<br>±1.45 | -3.28 | -4.22 – 2.33 | N.A. |
| Cylinder | -1.00<br>±1.02 | -2.44<br>±1.84 | 1.58 | 1.06 – 2.12 | N.A. | -0.93<br>±0.91 | -3.14<br>±1.86 | 2.21 | 1.39 – 3.03 | N.A. |
| Axis | 83<br>±45 | 92<br>±45 | -9 | -20 - 1 | N.A. | 81<br>±64 | 93<br>±20 | -53 | -143 - 37 | N.A. |
| CDVA LogMAR | 0.23<br>±0.35 | 0.00<br>±0.12 | 0.26 | 0.15 – 0.37 | <0.001 | 0.19<br>±0.27 | 0.01<br>±0.13 | 0.17 | 0.08 – 0.27 | 0.001 |
| CDVA Snellen | 0.72<br>±0.37 | 1.03<br>±0.28 | 0.38 | -0.51 - -0.27 | N.A. | 0.73<br>±0.32 | 1.01<br>±0.26 | -0.27 | -0.39 - -0.16 | N.A. |

Abbreviations: CDVA: corrected distance visual acuity, CI: confidence interval, D: diopter, , N.A.: not assessed, SEQ: spherical equivalent, logMAR: logarithm of the minimum angle of resolution for visual acuity
<sup>a</sup>Unless otherwise specified, reported as mean ±SD.
<sup>b</sup>Unless otherwise specified, reported as mean difference (web-based minus manifest assessment).
<sup>c</sup>Differences are based on the 54 and 26 cases with both manifest and digital refraction data available, leading to small deviations when subtracting the reported mean data in the table.
<sup>d</sup>Paired-sample Student t test was performed for predefined primary and secondary outcome parameters only.
<sup>e</sup>Spherical and cylindrical power and axes were translated into vectors using Fourier analysis and the difference is calculated as a power vector of the difference between the power vectors.
<sup>f</sup>The difference between power vectors and the vector specific parameters are calculated as a residual vector and is non-linear.

**Figure 3.**
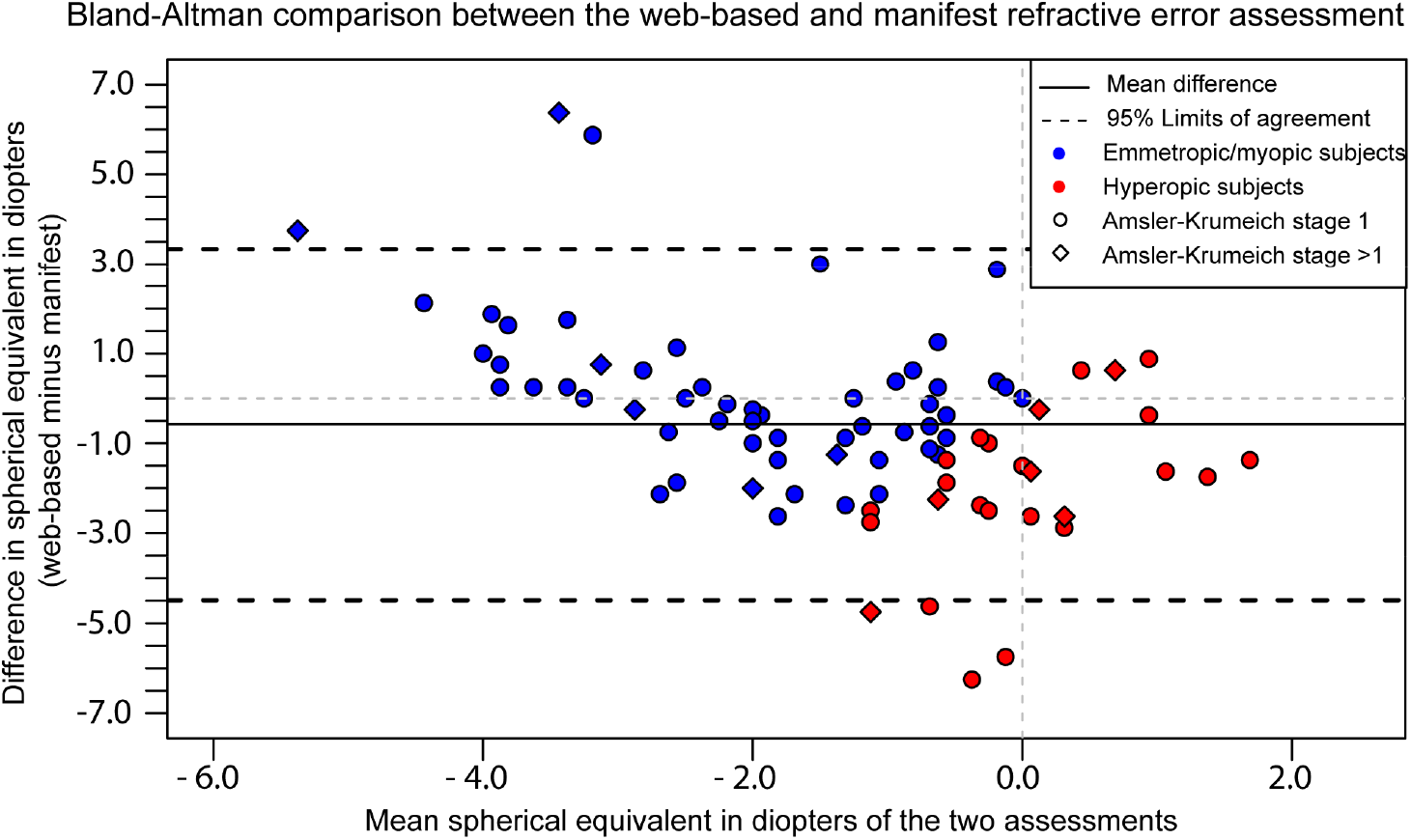
A Bland‐Altman plot displaying the differences in refractive error between the web‐based refractive assessment (index test) and the manifest refraction (reference test). The difference between the reference and index test shown on the Y‐axis is expressed as the difference of the web‐ based refractive assessment outcome compared to the manifest refraction. The x‐axis shows the mean spherical equivalent of the two assessments. Myopia and hyperopia were based on the spherical equivalent of the manifest refraction.

We performed a multivariable Generalized Estimating Equations (GEE) analysis to correct for the inclusion of two eyes of one patient, age, sex, and keratoconus severity (supplementary Table 3). As expected the Amsler‐Krumeich stage >1 (B=1.167, P=0.027) had a significant effect on the power vector of the manifest refraction, indicating the refractive error increases with keratoconus severity. The web‐based refraction did not identify this increase in power vector. Stratification of outcomes for myopia and hyperopia revealed no new insights.

The algorithm of the digital refraction was not always able to correctly determine the participant’s refractive error as either myopia (‐) or hyperopia (+). In a total of 21 cases (21%) the signation between the index test and the reference test differed, with an average absolute difference in refractive error of ‐2.38±1.96. Determining the correct signation in hyperopic subject proved challenging: in 20 of 27 (74%) of hyperopic subjects the signation switched between the index and reference test. Strikingly, only one myopic case was incorrectly identified by the online test (98% success). Naturally, this has profound effect on the attained corrected distance visual acuities withf the web‐based prescription.

### Corrected visual acuity measurement using the web‐based refraction measurement

The overall achieved corrected distance visual acuity was significantly lower with the digital prescription (0.22±0.32 logMAR) vs. the reference test (‐0.01±0.13 LogMAR, *P* <0.001). For myopic cases the mean difference in CDVA was 0.23 logMAR (95%CI ‐0.37 to ‐0.15; Snellen 0.31 95%CI 0.27 to 0.51). For hyperopic cases these outcomes were comparable (‐0.22logMAR; 95%CI ‐0.27 to ‐0.08; Snellen 0.31 95%CI 0.16 to 0.39). This underlines that the variation in refractive outcomes translate to visual acuity outcomes on average 3 lines less read on a visual acuity chart.

In 51 eyes (n=26 subjects) the CDVA was not assessed with the prescription of the web‐based refractive error assessment. In 13 eyes the web‐based refraction assessment did not yield an outcome because of the previous mentioned technical errors. In 19 consecutive assessed patients the CDVA was not assessed due to an incorrect instruction of a member of the research team. We identified no other relevant associations, and considered these data missing at random.

## Discussion

In this clinical method comparison study we compared a web‐based tool for measuring refractive errors with a manifest refraction, the current gold standard, in keratoconus patients. The relevance of delivering remote eye care has been illustrated during the COVID‐19 outbreak.(4) With an acute reduction in access to care during this period, the most relevant finding of our studies is that visual acuity can be assessed using a web‐based exam, in healthy subjects as well as in individuals with a complex refractive error and an ophthalmic condition.(6) In the high visual acuity group the agreement of the measurements is high. It should be noted that repeated visual acuity assessments always demonstrate variability due to measurement variation. Cross‐sectional studies on repeatability of clinical logMAR wallcharts revealed 95% limits of agreements of +‐ 0.15 logMAR.(21) When looking at the visualized differences between the web‐based test and the reference test we consider the agreement to be clinically acceptable. Notwithstanding, the spread in the outcomes of the web‐based refraction assessment exceeded the predefined non‐inferiority margins (of <0.5D), and poorly correlated with the gold standard manifest refraction (for myopia 95% LoA: ‐3.28 – 3.47, and for hyperopia 95% LoA: ‐5.52 – 1.39; ICC: 0.36). As a result, the attained CDVA with the prescription of the web‐based tool was significantly lower than the traditional manifest refraction (logMAR 0.23 vs. 0.00, P<0.001). It should be noted that the refractive assessments do not agree equally throughout the range of measurements. The agreement appears to improve in the low myopic refractive error range, suggesting a better performance of the web‐based refractive assessment in this range. Furthermore, we observed that the web‐based refractive assessment undercorrected higher myopics and overcorrected lower myopics. Both effects might by mitigated by a judicial re‐calibration of the algorithm of the online test.

The performance of the web‐based refractive assessment in this study population can be explained by the design of the algorithm. The algorithm translates the measured visual acuity in a refractive error. Next, the astigmatic refractive error is assessed and the spherical and cylindrical components are determined. The algorithm assumes that the loss of visual acuity is proportional to the increase in refractive error, and that all vision loss is caused by a refractive error. These two assumptions obviously do not necessarily stand in eyes with an ophthalmic condition.

The signation (either + or ‐) is assessed by a red/green test, and by asking the participant questions on the experience of their visual function (do you have problems reading, do you recognize faces from afar etc.). These questions appear to suffice in a healthy population (98% success)(6), but are not considerable reliable in this population (80% success): keratoconus patients often have problems with both near and far tasks, and the questions have not always been discriminative. This effect is more pronounced in hyperopic refractive errors and could be mitigated by feeding the algorithm more data than was available in the clinical trial, in particular data on any previous prescriptions. This is the case in the commercially available exam: an optometrist assesses any existing prescription and validates the findings that are produced by the algorithm. The here employed clinical trial algorithm functioned completely independent.

Several considerations of the study deserve attention. No randomization of the test order was performed, which could have impacted our results as subject may become tired during the assessments. However, because of the fixed test order, this should have impacted all subject similarly. The learning or training effect is considered negligible since the three refractive assessment methods are very different and randomized optotypes were used for assessment of the visual acuity. Observer bias cannot be excluded as the observer had access to all test results. A consideration should be made regarding the missing data. The primary outcome – refractive error‐ was missing in 13 eyes because of technical errors. The number of technical errors was evidently higher when compared to the healthy study population previously described (6/200 eyes).(6) The data were particularly missing in cases with higher refractive errors and lower visual acuities (i.e. the more severe cases), suggesting these cases are not missing at random. Apparently, the complex refractive errors pose a challenge for the algorithm. This may have impacted the study power. The missing data are considered to have little impact on our conclusions, since we already concluded that the algorithm’s performance is poor in this group. Lastly, the participants were young (25.6 years on average) and presumably digital natives: the uptake of these novel tools in an elderly population warrants a design tailored to their needs and digital aptitude.

The digital refraction fits into the current trend of health care digitalization as health care demand is increasing.(22) Importantly, this is expected to increase because of an aging population, whereas health care budgets are capped, and countries experience a shrinking workforce.(23–25) In addition, telemedicine has the ability to alleviate the urgent challenges our health care systems are currently facing. Several studies have shown the potential of digital eye care tools and the use of digital tools for diagnosis and monitoring of patients.(7,26–29) In particular, the PEEK vision tool developed by Bastawrous et al, provided health care professionals with accurate and vital information regarding a person’s ocular health status, and improved eye health in a rural African community.(7) Notwithstanding, our results show that validation is of upmost importance preceding clinical implementation of these tools and new iterations are needed to further improve the accuracy of the here studied web‐based assessment tool.

## Conclusions

The web‐based digital eye exam is able to deliver estimates of visual acuity, achieved independently and remotely by the patient. Contrary to the previously published results with healthy volunteers, the assessment of refractive error posed too big a challenge for the digital algorithm and its prescription resulted in a significantly poorer visual performance. These data will aid in a better training and calibration of the web‐based test, with the potential to better identify outlier measurements and increase overall test reliability. A web‐based exam should not be considered a replacement for a comprehensive manual examination by an eye care professional. Notwithstanding, the outcome of the digital eye exam can provide doctors with an objective measurement of visual function which enhances a teleconsultation. The latter is especially important in times or areas with limited access to health care.

## Data Availability

The data underlying the results are available upon request,
except for the uncorrected visual acuity data. The data can only be shared with other
researchers after signing a transfer agreement with the manufacturer of the tool,
easee.online.

## Abbreviations

AK: Amsler Krumeich
CDVA: Corrected distance visual acuity
CI: confidence interval
D: diopter
ETDRS: Early Treatment Diabetic Retinopathy Study
LoA: limits of agreement
LogMAR: logarithm of the minimum angle of resolution
GEE: generalized estimates equation
UDVA: uncorrected distance visual acuity
ICC: intraclass correlation coefficient

## Acknowledgements

The authors are grateful to Emma van Hamersveld BSc, for her assistance in composing the regulatory and ethical board review documentation, and to Nienke Soeters PhD, Veerle Berkhof BSc and Timon Legemaat BSc for their assistance in acquiring the data.

