## Supplemental Table 1 for "The evaluation of a web‐based tool for measuring the uncorrected visual acuity and refractive error in keratoconus eyes: a prospective open‐label method comparison study"

| Supplementary data table 1. The refractive error and visual acuity measured in emmetropic and myopic eyes stratified for keratoconus severity | | | | | | | | | | | | | | | | | | | | |
| --- | --- | --- | --- | --- | --- | --- | --- | --- | --- | --- | --- | --- | --- | --- | --- | --- | --- | --- | --- | --- |
|  | Total (n=71) | | | | | Amsler-Krumeich stage 1 (n=61) | | | | | Amsler-Krumeich stage 2 (n=8) | | | | | Amsler-Krumeich stage 3 (n=2) | | | | |
| Refractive error and visual acuity | Web-based refraction^a^ (n=54) | Manifest refraction^a^ (n=71) | Difference^b,c^ | 95% CI | P-value^d^ | Web-based refraction^a^ (n=48) | Manifest refraction^a^ (n=61) | Difference^b,c^ | 95% CI | P-value^d^ | Web-based refraction^a^ (n=5) | Manifest refraction^a^ (n=8) | Difference^b,c^ | 95% CI | P-value^d^ | Web-based refraction^a^ (n=1) | Manifest refraction^a^ (n=2) | Difference^b,c^ | 95% CI | P-value^d^ |
| Power vector ^e,f^ (D) | 2.34  ±1.02 | 2.69  ±1.74 | -1.08 | -1.41 – -0.74 | N.A. | 2.45 ±0.95 | 2.40 ±1.56 | -1.03 | -1.35 – -0.72 | N.A. | 1.93 ±1.09 | 4.04 ±1.53 | -1.40 | -3.95 –- 1.15 | N.A. | 1.80 | 6.31 ±1.81 | -1.13 | - | N.A. |
| Power Vector J0^f^ (X) | -0.08  ±0.54 | -0.61  ±1.17 | -1.16 | -1.41 – -0.90 | N.A. | -0.12 ±0.56 | -0.50 ±1.08 | -1.07 | -1.32 – -0.81 | N.A. | 0.11 ±0.13 | -1.02 ±1.63 | -1.46 | -2.66 –-0.26 | N.A. | - | -2.14 ±0.98 | - | - | N.A. |
| Power vector J45^f^ (Y) | 0.05  ±0.62 | 0.02  ±0.86 | -0.72 | -0.91 – -0.53 | N.A. | 0.05 ±0.66 | 0.147 ±0.81 | -0.75 | -0.17 –0.94 | N.A. | 0.03 ±0.23 | 0.31 ±1.10 | -0.57 | -1.27 –0.13 | N.A. | - | -1.11 ±0.87 | - | - | N.A. |
| SEQ (D) | -1.96  ±1.20 | -2.05  ±1.76 | 0.09 | -0.35 – 0.54 | 0.675 | -1.90 ±1.20 | -1.87 ±1.54 | -0.03 | -0.43 – 0.37 | 0.872 | -2.20 ±1.16 | -2.92 ±2.37 | 0.73 | -3.40 – 4.85 | 0.651 | -3.50 | -5.69 ±2.21 | - | - | - |
| Sphere | -1.53  ±1.15 | -0.87  ±1.79 | -0.76 | -1.23 – 0.28 | N.A. | -1.50 ±1.12 | -0.76 ±1.63 | -0.83 | -1.28 – 0.39 | N.A. | -1.45 ±1.23 | -1.28 ±2.52 | -0.35 | -4.90 – 4.20 | N.A. | -3.50 | -3.13 ±2.65 | - | - | N.A. |
| Cylinder | -1.00  ±1.02 | -2.44  ±1.84 | 1.58 | 1.06 – 2.12 | N.A. | -0.95 ±0.99 | -2.30 ±1.64 | 1.53 | 0.97 – 2.09 | N.A. | -1.50 ±1.27 | -3.88 ±1.88 | 2.15 | -0.31 –  4.61 | N.A. | - | -5.13 ±0.88 | - | - | N.A. |
| Axis | 83  ±45 | 92  ±45 | -9 | -20 - 1 | N.A. | 82  ±43 | 90 ±42 | -9 | -20 - 2 | N.A. | 91  ±65 | 98  ±38 | -13 | -65  -  38 | N.A. | - | 105 ±14 | - | - | N.A. |
| CDVA logMAR | 0.23  ±0.35 | 0.00  ±0.12 | 0.26 | 0.15 – 0.37 | <0.001 | 0.18 ±0.28 | -0.02 ±0.11 | 0.22 | 0.12 –  0.32 | <0.001 | 0.80 | 0.09 ±0.10 | - | - | N.A. | 1.30 | 0.29 ±0.15 | - | - | N.A. |
| CDVA snellen | 0.72  ±0.37 | 1.03  ±0.28 | 0.38 | -0.51 - -0.27 | N.A. | 0.76 ±0.34 | 1.07 ±0.26 | -0.39 | -0.51 - -0.27 | N.A. | 0.16 | 0.83 ±0.19 | - | - | N.A. | 0.05 | 0.53 ±0.18 | - | - | N.A. |
| Notes: Abbreviations: CDVA: corrected distance visual acuity, N.A.: not assessed, SEQ: spherical equivalent, logMAR: logarithm of the minimum angle of resolution for visual acuity ^a^ Unless otherwise specified, reported as mean (SD). ^b^ Unless otherwise specified, reported as mean difference (web-based minus manifest assessment).  ^c^ Paired-sample Student t test was performed for predefined primary and secondary outcome parameters only.  ^d^ Differences are based on the cases with both manifest and web-based refraction data available, leading to small deviations when subtracting the reported mean data in the table. ^e^ Spherical and cylindrical power and axes were translated into vectors using Fourier analysis and the difference is calculated as a power vector of the difference between the power vectors. ^f^ The difference between power vectors and the vector specific parameters are calculated as a residual vector and is non-linear. | | | | | | | | | | | | | | | | | | | | |
