## Supplemental Table 2 for "The evaluation of a web‐based tool for measuring the uncorrected visual acuity and refractive error in keratoconus eyes: a prospective open‐label method comparison study"

| Supplementary data table 2. The refractive error and visual acuity measured in hyperopic eyes stratified for keratoconus severity | | | | | | | | | | | | | | | | | | | | |
| --- | --- | --- | --- | --- | --- | --- | --- | --- | --- | --- | --- | --- | --- | --- | --- | --- | --- | --- | --- | --- |
|  | Total (n=29) | | | | | Amsler-Krumeich stage 1 (n=23) | | | | | Amsler-Krumeich stage 2 (n=5) | | | | | Amsler-Krumeich stage 3 (n=1) | | | | |
| Refractive error and visual acuity | Web-based refraction^a^ (n=54) | Manifest refraction^a^ (n=71) | Difference^b,c^ | 95% CI | P-value^d^ | Web-based refraction^a^ (n=48) | Manifest refraction^a^ (n=61) | Difference^b,c^ | 95% CI | P-value^d^ | Web-based refraction^a^ (n=5) | Manifest refraction^a^ (n=8) | Difference^b,c^ | 95% CI | P-value^d^ | Web-based refraction^a^ (n=1) | Manifest refraction^a^ (n=2) | Difference^b,c^ | 95% CI | P-value^d^ |
| Power vector^e,f^ (D) | 1.35  ±0.68 | 2.02  ±1.04 | -0.69 | -0.92 – -0.46 | N.A. | 1.44 ±0.67 | 2.10 ±1.05 | -0.59 | -0.81 –-0.38 | N.A. | 1.03 ±0.40 | 1.84 ±1.07 | -1.00 | - | N.A. | 1.78 | 0.90 | -1.82 | - | N.A. |
| Power Vector J0^f^ (X) | -0.08  ±0.26 | -1.27  ±1.01 | -1.47 | -2.00 – -0.93 | N.A. | 0.13 ±0.25 | -1.28 ±1.08 | -1.38 | -1.97 –-0.78 | N.A. | 0.24 ±0.01 | -1.50 ±0.69 | -2.13 | - | N.A. | 0.24 | -0.57 | - | - | N.A. |
| Power vector J45^f^ (Y) | -0.04  ±0.23 | -0.16  ±1.00 | -0.98 | -1.41 – -0.54 | N.A. | -0.04 ±0.23 | -0.10 ±0.99 | -0.88 | -1.29 –-0.47 | N.A. | 0.08 | -0.43 ±1.32 | -2.38 | - | N.A. | 0.24 | -0.48 | - | - | N.A. |
| SEQ (D) | -1.02  ±1.40 | 1.04  ±0.83 | -2.06 | -2.76 – -1.37 | <0.001 | -1.02 ±1.39 | 1.11 ±0.87 | -2.14 | -2.94 – -1.33 | <0.001 | -0.85 ±1.85 | 0.88 ±0.58 | -1.73 | -4.33 – 0.89 | 0.140 | -1.75 | 0.50 | - | - | - |
| Sphere | -0.59  ±1.39 | 2.68  ±1.45 | -3.28 | -4.22 – 2.33 | N.A. | -0.56 ±1.44 | 2.83 ±1.48 | -3.39 | -4.51 - -2.28 | N.A. | -0.55 ±1.45 | 2.35 ±1.43 | -2.90 | -6.16 – 0.36 | N.A. | -1.50 | 1.25 | - | - | N.A. |
| Cylinder | -0.93  ±0.91 | -3.14  ±1.86 | 2.21 | 1.39 – 3.03 | N.A. | -1.04 ±0.99 | -3.27 ±1.80 | 2.24 | 1.22 – 3.25 | N.A. | -0.60 ±0.55 | -2.95 ±2.35 | 2.35 | 0.00 – 4.70 | N.A. | -0.50 | -1.50 | - | - | N.A. |
| Axis | 81  ±64 | 93  ±20 | -53 | -143 - 37 | N.A. | 76 ±66 | 91  ±21 | -67 | -183 - 49 | N.A. | 78 ±48 | 98 ±18 | -20 | -105 - 66 | N.A. | 171 | 110 | - | - | N.A. |
| CDVA logMAR | 0.19  ±0.27 | 0.01  ±0.13 | 0.17 | 0.08 – 0.27 | 0.001 | 0.24 ±0.30 | 0.01 ±0.13 | 0.22 | 0.10 – 0.35 | 0.003 | 0.08 ±0.17 | 0.02 ±0.13 | 0.06 | -0.03 – 0.15 | 0.149 | - | 0.07 | - | - | N.A. |
| CDVA snellen | 0.73  ±0.32 | 1.01  ±0.26 | -0.27 | -0.39 - -0.16 | N.A. | 0.66 ±0.25 | 1.02 ±0.26 | -0.36 | -0.49 - -0.23 | N.A. | 0.89 ±0.42 | 0.98 ±0.31 | -0.09 | -0.29 – 0.10 | N.A. | - | 0.85 | - | - | N.A. |
| Notes:  CDVA: corrected distance visual acuity, N.A.: not assessed, SEQ: spherical equivalent, logMAR: logarithm of the minimum angle of resolution for visual acuity  ^a^ Unless otherwise specified, reported as mean (SD).  ^b^ Unless otherwise specified, reported as mean difference (web-based minus manifest assessment).  ^c^ Differences are based on the cases with both manifest and web-based refraction data available, leading to small deviations when subtracting the reported mean data in the table.  ^d^ Paired-sample Student t test was performed for predefined primary and secondary outcome parameters only.  ^e^ Spherical and cylindrical power and axes were translated into vectors using Fourier analysis and the difference is calculated as a power vector of the difference between the power vectors.  ^f^ The difference between power vectors and the vector specific parameters are calculated as a residual vector and is non-linear. | | | | | | | | | | | | | | | | | | | | |

Supplementary data table 2. The refractive error and visual acuity measured in hyperopic eyes stratified for keratoconus severity
