## Supplemental Table 3 for "The evaluation of a web‐based tool for measuring the uncorrected visual acuity and refractive error in keratoconus eyes: a prospective open‐label method comparison study"

| Supplementary table 3. Multivariate analysis to identify the associations between independent variables and the refractive error outcome for the difference between the web-based refractive assessment and manifest refraction, the web-based refractive assessment and the manifest refraction. | | | | | | | | | |
| --- | --- | --- | --- | --- | --- | --- | --- | --- | --- |
|  | Dioptric difference between the power vectors of the web-based and manifest refractive assessments ^a^ | | | Power vector web-based refractive assessment | | | Power vector manifest refraction | | |
|  | B | 95% Confidence interval | P-value^b^ | B | 95% Confidence interval | P-value^b^ | B | 95% Confidence interval | P-value^b^ |
| Sex^c^ | 0.611 | -0.172 to 1.396 | 0.126 | 0.187 | -0.583 to 0.958 | 0.634 | -0.206 | -1.048 to 0.635 | 0.631 |
| Age | 0.062 | -0.003 to 0.128 | 0.61 | 0.057 | 0.000 to 0.115 | 0.052 | 0.026 | -0.046 to 0.097 | 0.485 |
| Amsler-Krumeich stage >1^d^ | 0.889 | -0.195 to 1.972 | 0.108 | 0.127 | -0.585 to 0.839 | 0.726 | 1.167 | 0.132 to 2.202 | 0.027 |
| B; beta value  ^a^ Absolute difference between power vectors calculated as an residual vector (web-based minus manifest)  ^b^ Analysed using an Generalized Estimating Equations to correct for inclusion of 2 eyes of one patient  ^c^ Reference male  ^d^ Reference Amsler-Krumeich stage 1 | | | | | | | | | |
